# ‘Ready for tomorrow: eHealth readiness in top clinical hospitals surveyed’

**DOI:** 10.64898/2025.12.22.25342805

**Authors:** G.A.G. Garritsen, M.E.M. den Ouden, N. Beerlage-de Jong, S. M. Kelders

**Author notes:** **Corresponding Author:** G. A.G. Garritsen, Research Group Technology, Health & Care Research Group, Saxion University of Applied Sciences, M.H. Tromplaan 28 Enschede, 7513 AB Netherlands, | (GG).

## Abstract

Technological innovations such as eHealth are vital for improving healthcare accessibility, quality, and sustainability. While most research addresses adoption at the individual or team level, less is known about organisational factors enabling sustainable transformation. Organisational readiness is a key determinant of success. The Organizational eHealth Readiness (OeHR) model, developed in Polish primary care, assesses five dimensions: Strategy, Competence, Culture, Structure, and Technology, but its applicability in Dutch health care remains unclear.

This mixed-methods study evaluated the OeHR model in Dutch hospitals. A validated 32-item questionnaire, translated into Dutch, was completed by managers and implementation specialists of 15 top-clinical hospitals (n=22). Descriptive statistics, regression analyses, and Principal Component Analysis provided insight into how the five dimensions jointly reflect organisational readiness for eHealth. Three focus groups (n=14) in two hospitals explored construct interpretation, missing dimensions, and model usability. Qualitative data were analysed using deductive coding on OeHR dimensions and emergent themes to refine the questionnaire.

Quantitative analyses identified organisational culture as the only significant predictor of subjective eHealth readiness, while other dimensions showed no independent effect. Open responses and focus groups confirmed the centrality of culture and suggested refinements to all components, including clearer definitions, structural flexibility, and attention to external factors. Overall, the OeHR model was valued for strategic guidance but required contextualisation for practical use.

This study showed that the OeHR model provides a valuable framework for assessing eHealth readiness in Dutch hospitals, with cultural readiness emerging as the most influential yet conceptually ambiguous dimension. Strategy, competence, structural and technological readiness mainly act as contextual enablers rather than direct predictors. The findings highlight the need to refine definitions, reduce overlap, and explore additional layers such as personal, operational, and societal readiness. Strengthening conceptual clarity and developing context-sensitive tools could enhance applicability and guide hospitals in translating readiness into digital transformation.

**Author summary:** eHealth, such as home monitoring and online consultations with healthcare providers, is becoming increasingly important. Hospitals want to implement eHealth effectively, but this is only possible if the organisation is ready for it. The Organisational eHealth Readiness model helps to assess how “eHealth-ready” an organisation is. This model looks at five components: strategy, competencies, culture, structure and technology.

In our research, we examined whether this model is also suitable for Dutch hospitals and which factors were still missing. We did this using a questionnaire among professionals and three focus groups, all involved in the implementation of eHealth. This showed that all components of the model are important, but that the culture of the organisation plays a central role. If employees are open to change and innovation, this acts as a driving force for eHealth. The other components, such as technology and strategy, appear to be primarily preconditions: necessary, but not sufficient to enable real change.

Participants found the model useful, but felt that some factors were missing, such as leadership, collaboration, flexibility and the influence of legislation and regulations. They also mentioned that the organisation must have the capacity to cope with change effectively. Adapting the model to include these points could better support hospitals in healthcare transformation in the future.

## Introduction

Technological innovations are essential to increase the accessibility, quality, and sustainability of healthcare systems [1]. One promising innovation is eHealth, defined as the use of digital information and communication to support health and care [1]. In hospitals, eHealth can improve processes and quality of care through better communication and collaboration between providers and patients [2]. This can also help shift care from the hospital to the homes of people. Home monitoring exemplifies this shift, enabling patient care outside the hospital and transforming reactive care into proactive practice [3].

However, such a shift requires profound changes in the behaviour of healthcare professionals and in the structure of healthcare organisations. For example, collaboration across disciplines and domains is crucial, yet often difficult to achieve in practice [4,5]. To date, research on the implementation of eHealth has primarily focused on the adoption and implementation of technologies at the individual or team level, particularly examining factors that influence user acceptance, usability, and integration into daily routines of end-users such as healthcare professionals and patients [6–8].

Key insights from previous studies highlight the importance of digital skills, innovation-promoting cultures, and effective communication during change processes. Consequently, we lack a broader understanding of the organisational dynamics that fundamentally shape the success or failure of such transitions. This gap is especially pressing given the current absence of validated, practice-based tools that can strategically support organisations in transforming care [2].

After all, individuals always operate within an organisational context. This context has a significant impact on the sustainability and scalability of eHealth implementations [9–11]. A more integrated perspective that also incorporates an organisation’s readiness for implementing eHealth is therefore essential to understand and support digital healthcare innovation.

When considering readiness, it is important to distinguish between individual and organisational change readiness. Individual change readiness relates to personal skills, while organisational change readiness concerns the capabilities and structures of the organisation [12]. Weiner (2009) defines organisational change readiness as “the extent to which an organisation is prepared for a planned change, determined by the needs and expectations from the environment” [11].

To assess such preparedness in the context of digital health, Kruszyńska-Fischbach et al. (2022) developed the Organizational eHealth Readiness (OeHR) model. Originating in Polish primary care, the model integrates dimensions of readiness for change with five key organisational dimensions:

- Strategic eHealth readiness: the extent to which digital innovation is embedded in strategic objectives [13,18]
- Competence eHealth readiness: the deployment of resources and training for digitally supportive, patient-centred work [14,18]
- Cultural eHealth readiness: the degree to which culture and norms within the organisation support digital care [15,18]
- Structural eHealth readiness: the presence of structures, policies, leadership, and processes for eHealth adoption [15,16,18]
- Technological eHealth readiness: the availability and quality of infrastructure, technical expertise, and ICT support [15,17,18]

The OeHR model has proven useful for assessing eHealth readiness in Polish primary care [18]. However, its relevance as a diagnostic tool to anticipate barriers and guide improvement in other settings, in our case Dutch secondary care - remains unclear. The study aims to explore how stakeholders in Dutch hospitals perceive and evaluate the OeHR model, and to examine quantitatively how its components relate to the perceived overall eHealth readiness of their organisations. This mixed-methods approach provides a more comprehensive view of organisational eHealth readiness and its underlying dimensions.

## Method

### Ethic statement

This study was approved by the Ethics Committee of the University of Twente (reference number 231274). All participants gave informed consent for the survey and participating in the focus group.

### Design of the study

A mixed-methods exploratory design was used to investigate the relevance of the OeHR model for the Dutch hospital setting. The study focused both on the perceived relevance of the existing OeHR dimensions and on identification of potential missing factors. First, quantitative data were collected using a structured questionnaire based on the original OeHR model. This provided insight into how innovation managers, programme managers and healthcare professionals involved in eHealth implementations within Dutch hospitals evaluated each dimension. Additionally, to deepen this understanding and to explore potentially overlooked items and dimensions, focus groups were conducted.

### Participants

#### Questionnaire

Participants were recruited from 15 top clinical hospitals (STZ hospitals) across the Netherlands through two hospital partnerships (Santeon and mProve). These hospitals are characterised by their combination of patient care, clinical training and research into complex conditions and often serve a supra-regional population [19]. Within these hospitals, we recruited individuals who play an essential role in digital transformation processes because of their direct involvement in patient care, technological innovation, strategic planning and/or operational implementation. Therefore, healthcare professionals with eHealth implementation experience, programme, innovation and ICT managers were invited to participate.

#### Focus groups

Two recruitment strategies were used to invite participants for the focus groups. First, at the end of the questionnaire, respondents were asked via a yes/no question whether they were interested in taking part in a follow-up focus group. Those who were interested were subsequently invited. Second, additional participants were recruited through two regional hospitals, where the focus groups were integrated into existing scheduled meetings. In total, three focus groups were conducted across two hospitals, involving 14 participants. Two focus groups were held at Medisch Spectrum Twente (MST; labelled MST 1 and MST 2 in the results), and one focus group at Zorggroep Twente (ZGT; labelled ZGT 1 in the results). The same inclusion criteria as were applied for the focus group and questionnaire. Hence, participants included innovation and programme managers, ICT managers, and healthcare professionals involved in the implementation of eHealth and value-based healthcare.

### Materials

#### Questionnaire

The validated questionnaire on organisational eHealth readiness developed by Kruszyńska-Fischbach et al. (2022) was used. The original instrument consists of 32 items across five dimensions, each capturing a key aspect of organisational readiness: Strategic eHealth readiness (n = 4), competence eHealth readiness (n = 6), cultural eHealth readiness (n = 10), structural eHealth readiness (n = 7), and technological eHealth readiness (n = 5) [18]. Items address, for example, the extent to which eHealth is embedded in strategic policy. They also cover the level of digital skills among professionals and the openness of organisational culture to innovation. In addition, the questionnaire includes items on the availability of supportive structures and resources. Finally, several items assess the maturity of technological infrastructure. All items were rated on a 5-point Likert scale ranging from “Totally disagree” to “Totally agree”. In line with World Health Organisation (WHO) guidelines for instrument translation and adaptation [20], the questionnaire was translated from English to Dutch using a forward–backward translation process. A team of three bilingual experts, including one with healthcare experience, independently translated the questionnaire into Dutch, after which a consensus version was developed. A fourth independent expert then conducted a back-translation into English to verify content accuracy. Following comparison and minor adjustments, the preliminary Dutch version was tested using the “think-aloud” method [21] with a digital transformation programme manager to identify ambiguities or interpretation issues. This led to small wording changes to ensure contextual relevance for a hospital setting.

Next to the validated OeHR questionnaire, we added several open-ended items to gain additional insight into the perceived applicability and completeness of the instrument. These items assessed: (1) whether the components of the OeHR questionnaire were considered clear and complete; (2) whether additional topics or components relevant to organisational eHealth readiness were missing; (3) how respondents relate OeHR to broader healthcare transformation processes; and (4) experiences with the implementation and up-scaling of eHealth solutions. Lastly, the perceived (subjective) Organisational eHealth Readiness (OeHR) was assessed by asking participants to rate their organisations overall perceived Organisation eHealth Readiness on a scale from 1-10.

#### Focus groups

The focus groups aimed to: (1) assess the applicability and relevance of the OeHR model and questionnaire in the Dutch hospital context, (2) identify potentially missing items or dimensions within the model, and (3) explore how the OeHR model could support care transformation within healthcare organisations. A structured discussion guide was used during the focus groups to ensure comparability across focus groups. The guide included an introduction of the study objectives and an explanation of the OeHR model, including its five dimensions. Each focus group explored participants’ views on the relevance and interpretation of the five OeHR dimensions: strategy, competence, culture, structure, and technology. To guide the discussion, participants were asked descriptive questions (e.g., “How is this aspect of eHealth readiness currently addressed in your organisation?”) and evaluative questions (e.g., “To what extent do you consider this dimension relevant or well developed in your hospital?”). In addition, participants were explicitly invited to reflect on the completeness of the model and questionnaire, for example by discussing whether they felt any dimensions or items were missing or insufficiently represented. Finally, reflective and generative prompts were used to explore how the OeHR model could be further improved or applied in practice (e.g., “How might this model support your organisation’s digital transformation efforts?”).

### Procedures

#### Questionnaire

Potential participants received an information letter explaining the aim and procedure of the study and a link to the online questionnaire (in Qualtrics) [35]. Participation was voluntary and informed consent (digitally) signed before the start of the questionnaire. The survey was completed anonymously; participants’ data could not be traced back to individuals. If participants provided contact details for participation in follow-up focus groups, these data were stored separately to ensure confidentiality.

#### Focus groups

Two researchers led the focus group discussions, and the use of a shared discussion guide ensured a consistent process and comparability across focus groups. The focus groups, held in March and April 2025, lasted approximately one hour. Each began with an explanation of the study’s aim and a brief presentation of the OeHR model. Participants reflected on the dimensions using interactive methods such as sticker voting, post-it notes, and group discussion, to assess the perceived relevance of the dimensions and identify potentially missing items. In a second round, they discussed the practical applicability of the OeHR model in their own organisational setting. All sessions were audio-recorded and supplemented with photographs of flipchart materials.

To support the qualitative analysis, participants were also asked to prioritise keywords derived from the items within each dimension that they considered most relevant. This ranking provided deeper insight into which items participants regarded as most meaningful for their organisational context. The five dimensions were subsequently used as overarching thematic codes during the data-analysis.

### Data analysis

#### Questionnaire

Descriptive statistics (median, interquartile range and standardised z-scores) were calculated for all dimensions of the Organisational eHealth Readiness (OeHR) instrument to summarise the distribution of responses. The five OeHR dimensions were calculated as mean scores based on items on a 5-point Likert scale. To examine the underlying structure of the OeHR instrument and to assess whether the item structure corresponded to the theoretically proposed framework, a PCA was performed on the scores of the five dimensions. The analyses were performed on data from 22 respondents (N = 22), and the principal component analysis (PCA) included five variables (p = 5) corresponding to the five OeHR dimensions. Prior to this, the suitability of the data for PCA was evaluated using the Kaiser-Meyer-Olkin (KMO) and Bartlett’s sphericity test to ensure that the correlation matrix met the assumptions required for component extraction. PCA was used to identify the shared variance between the dimensions, investigate possible overlaps, and determine whether the dimensions represented empirically distinct or uniform constructs. Component extraction followed the Kaiser criterion (eigenvalues > 1), and communalities and component loadings were inspected to evaluate the contribution of each dimension to the component structure. Because the theoretical model assumes that the five OeHR dimensions are related and may exhibit considerable overlap, multicollinearity was assessed using Variance Inflation Factors (VIF) within an initial multiple regression model in which the five dimensions were entered as predictors of overall perceived organisational eHealth readiness (on a scale from 1-10). The presence of substantial multicollinearity informed the decision to rely on PCA-derived component scores in subsequent regression analyses. These component scores were used as predictors in a follow-up regression model to examine the association between the consolidated readiness construct and the participants’s overall perceived organisational eHealth readiness, thereby ensuring that collinearity did not bias the estimation of regression coefficients. All analyses were carried out using IBM SPSS Statistics (version 29).

Additionally, the open-ended questions on the appropriateness, clarity, and completeness of the items were analysed using a qualitative coding process. This was done for each of the five dimensions of the OeHR model. Using Microsoft Excel software, the data were coded deductively, applying predefined categories derived from the dimensions of the OeHR model.

#### Focus groups

The analysis of the focus group data was conducted according to the method of qualitative content analysis [23]. Transcripts of the three focus groups were transcribed verbatim and read several times to gain a thorough understanding of the content and to identify patterns related to consensus, prioritisation, and construct interpretation. In the organising phase, all data were coded in Atlas.ti 8 using a structured codebook based on the five dimensions of the OeHR model. Within these main dimensions, subcodes were created inductively to capture more detailed aspects of the discussions. Specifically, four types of data were analysed: (1) individual prioritisation of dimensions, which was qualitatively examined to identify shared patterns and divergences; (2) remarks about the relevance and suitability of different items; (3) statements regarding potentially missing items within dimensions; and (4) reflections on the overall applicability of the OeHR model, which were categorised inductively.

A second reviewer (MdO) was involved in the coding process to ensure reliability and consistency. This reviewer did not independently code the data but checked the coding conducted by the primary researcher to verify its correctness. The analysis was performed by one independent researcher.

## Results

### Questionnaire

#### Reflection on existing OeHR

In total, 30 respondents filled out the OeHR-questionnaire, of which 21 respondents completed the entire questionnaire, and one respondent completed more than 70%. The responses of the one participant who completed 70% of the questionnaire were retained for analysis, as instruments with this level of completeness are generally considered reliable for inclusion [34] Hence, data of 22 respondents were included in the present study. Table 1 provides an overview of the main results of the quantitative analysis per dimension of eHealth Readiness.

**Table 1.** Descriptive Statistics for Organisational eHealth Readiness Dimensions.

| Dimension | Median | IQR | $\alpha$ | Z |
| --- | --- | --- | --- | --- |
| Strategic eHealth Readiness | 4.3 | 0.8 | 0.73 | -1.58 |
| eHealth Competence Readiness | 3.6 | 0.5 | 0.69 | -0.06 |
| Cultural eHealth Readiness | 3.7 | 0.9 | 0.91 | 3.19 |
| Structural eHealth Readiness | 3.6 | 0.3 | 0.69 | -0.71 |
| Technological eHealth Readiness | 3.4 | 0.6 | 0.57 | -0.93 |

Median = median score per dimension; IQR = interquartile range; $\alpha$ = Cronbach's alpha; Z = z-score. Model statistics: $R^2 = .783$ , Model $p = .0001$ .

Internal consistency of the five OeHR dimensions was acceptable, with Cronbach’s alpha values ranging from .60 to .90, indicating sufficient reliability for exploratory analysis. Descriptive statistics showed that Strategic eHealth Readiness had the highest central tendency (Median = 4.3, IQR = 0.8), suggesting that care transformation is relatively well embedded at the strategic level of the participating organisations. eHealth Competence Readiness (Median = 3.6, IQR = 0.5) and Cultural eHealth Readiness (Median = 3.7, IQR = 0.9) showed moderately high central scores, while Structural (Median = 3.6, IQR = 0.3) and Technological eHealth Readiness (Median = 3.4, IQR = 0.6) were rated slightly lower. The z-scores indicate the relative dispersion of each dimension around the overall mean: Cultural eHealth Readiness showed the greatest positive deviation (z = 3.2), while the other dimensions exhibited minor or negative deviations, suggesting less influence or lower relative prominence in respondents’ perceptions.

Collinearity diagnostics indicated substantial multicollinearity among the OeHR predictors, particularly for Cultural Readiness (Tolerance = .071; VIF = 14.01). To address this issue, a Principal Component Analysis (PCA) was conducted. The PCA confirmed that the five readiness dimensions were strongly interrelated, resulting in one dominant component (KMO = .675; Bartlett’s χ²(10) = 66.918, p < .001). As shown in Table 2, the first component had an eigenvalue of 3.332 and accounted for 66.64% of the total variance, with all subsequent components having eigenvalues well below 1. This indicates that only the first component met the Kaiser criterion and thus represents a unidimensional underlying structure.

**Table 2.**
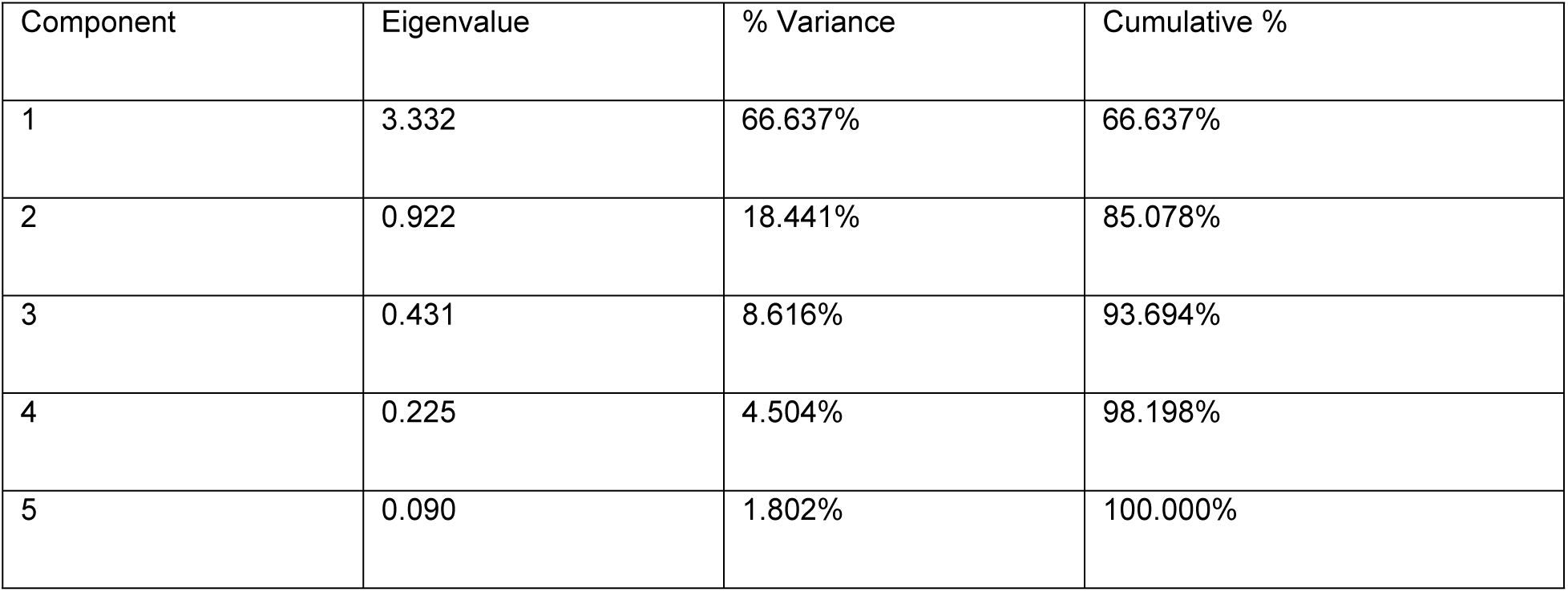
Explained Variance.

As shown in Table 3, the communalities indicate that Cultural eHealth Readiness has a particularly high extraction value (.854), suggesting that this dimension strongly captures the shared variance among the readiness factors and is therefore highly representative of the underlying construct.

**Table 3.** Communalities of the OeHR.

| Dimension | Initial | Extraction |
| --- | --- | --- |
| Strategic eHealth readiness | 1.000 | 0.649 |
| eHealth competence readiness | 1.000 | 0.737 |
| Cultural eHealth readiness | 1.000 | 0.854 |
| Structural eHealth readiness | 1.000 | 0.665 |
| Technical eHealth readiness | 1.000 | 0.428 |

In addition, the component loadings presented in Table 4 demonstrate that all five dimensions loaded substantially on the first component (ranging from .654 to .924), providing further evidence that they represent a single underlying construct.

**Table 4.** Component loadings per dimension of OeHR.

| Dimension | Loading |
| --- | --- |
| Strategic eHealth readiness | 0.805 |
| eHealth competence readiness | 0.858 |
| Cultural eHealth readiness | 0.924 |
| Structural eHealth readiness | 0.816 |
| Technical eHealth readiness | 0.654 |

The component score derived from this PCA was subsequently used in a regression analysis, demonstrating that the composite OeHR dimension significantly predicted perceived organizational eHealth readiness (β = .821, p < .001; R² = .674). These findings suggest that the five dimensions together reflect a single, integrated readiness construct, with cultural aspects playing a particularly pivotal role.

#### Missing items and dimensions

The results of the open questions showed that 91% of respondents considered the items suitable for assessing the five dimensions of organisational e-health readiness (OeHR). Of the respondents, 19% noted that items were missing or that the definitions were insufficient/unclear. They identified, opportunities to improve the conceptual clarity and completeness of the model. For example, some respondents indicated that strategic eHealth readiness should not only involve the Executive Board but also the tactical and operational management layers. Others reflected on the clarity of terms such as digital skills *‘Which category of digital skills do we mean?’ (Participant* 3) and noted that the term technological readiness may be too broad or vague “*Ready for what*?’ (*Participant 2).* Table 5 below summarises the number and nature of the suggestions made per dimension:

**Table 5.** Summary of suggested additions and revisions to the OeHR Model by dimension.

| OeHR Dimension | Suggested adjustment/addition | n (respondents) |
| --- | --- | --- |
| Strategic eHealth<br>Readiness | Vision and policy integration (embedding eHealth in annual plans, long-term strategies, and explicit strategic goals) | 2 |
|  | Sustainable funding and business case development (structural budgeting, not relying solely on temporary subsidies) | 2 |
|  | Governance and broad organisational support (beyond the Board of Directors, including management layers) | 1 |
|  | Innovative and developmental capacity (ability of the organisation to develop or stimulate eHealth solutions internally) | 1 |
| Competence<br>eHealth<br>Readiness | Digital competences of patients/clients | 1 |
|  | Differences in skills between organisational levels | 1 |
|  | Lack of structural support (e.g. training and job profiles) for competence development | 1 |
| Cultural eHealth<br>Readiness | Employee engagement, change readiness, and broad organisational commitment | 2 |
|  | Digital inclusion and accessibility for all users | 1 |
|  | Cultural support for adoption process | 1 |
|  | Focus on employee wellbeing (workload reduction, work satisfaction) | 1 |
| Structural eHealth<br>Readiness | Time and space for healthcare professionals to effectively and efficiently implement eHealth | 2 |
| Technological<br>eHealth<br>Readiness | Distinction between phases of technological development (technological in shape → optimisation → transformation) | 1 |

In addition, one respondent suggested adding additional dimensions, including personal e-health readiness (individual skills and readiness), operational e-health readiness (feasibility and support for e-health initiatives) and societal e-health readiness (societal readiness, including legal and financing aspects).

#### Overall applicability of the OeHR

Participants generally regarded the OeHR questionnaire as a valuable tool for reflection on eHealth Readiness. It helped raise awareness and introduced new perspectives on organisational readiness. According to participants, the OeHR questionnaire was most valuable (or applicable) for management or as input prior to major eHealth implementations projects. One participant commented, *‘It would be a great way to identify prior to a major e-health implementation, which should lead to care transformation, where the focus should be’. (participant 1).* Less practical applicability was foreseen for professionals in operational roles.

### Focus groups

The respondents from the three focus groups categorised the five dimensions of OeHR independently on relevance (1 = least relevance and 5 = most relevance). Table 6 shows that cultural eHealth readiness was rated as most relevant (median 4.5), while structural readiness was considered least relevant (median 2.0). The other dimensions received moderate relevance scores (median 3.0). Interquartile ranges (IQRs) revealed differences in the level of agreement: cultural and competence readiness showed high consensus (IQR 1.0), while strategic and structural readiness showed larger variation (IQR 2.5). These scores indicate varying perceived importance across the five OeHR dimensions.

**Table 6.** Median and Inter Quartile Range of the relevance of the OeHR dimensions.

| Dimensions OeHR | Median | IQR |
| --- | --- | --- |
| Strategic eHealth Readiness | 3,0 | 2,5 |
| eHealth Competence<br>Readiness | 3,0 | 1 |
| Cultural eHealth Readiness | 4,5 | 1 |
| Structural eHealth Readiness | 2,0 | 2,5 |
| Technological eHealth<br>Readiness | 3,0 | 2 |

The qualitative analysis of the three focus groups revealed clear patterns in the way participants interpreted and prioritised the five OeHR dimensions. Appendix 1 lists the prioritised keywords for each dimension.

Strategic readiness was consistently regarded as essential, provided that strategic choices aligned with concrete healthcare goals and daily practice. Competence readiness was strongly associated with digital skills, training, and leadership. Digital competence was seen as a fundamental prerequisite: *“You can build great systems, but if people can’t use them, they’re worthless”*. (ZGT 1)

Cultural readiness received high prioritisation, though interpretations varied. Themes such as change readiness, ownership, innovation culture, and communication were emphasised. Behavioural willingness to change was seen as crucial: *“If people are not willing to work differently, then you can structure what you want”*. (MST 1)

Structural readiness received relatively low prioritisation, although participants acknowledged the importance of standardisation and collaboration: *“Without standardisation from processes, every new application is doomed to fail”.* (ZGT 2)

Technological readiness was broadly recognised as a prerequisite. Reliable IT infrastructure was considered essential: *“First get the basics right, only then pursue major innovations”.* (MST 2) Participants also stressed the importance of involving end-users in co-development.

#### Missing items

The focus groups also highlighted items absent from the current OeHR model. Within the structural dimension, participants stressed the need for greater flexibility: *“Many processes are focused on control, whereas we are now looking for flexibility”.* (ZGT 1) A further structural gap concerned the lack of integration between daily care (“run”) and structural innovation (“change”): *“Change is about the innovations we develop and eventually have to integrate into the existing organisation*” (MST 2). Collaboration, both internal and external, was likewise identified as crucial for structural readiness but seen as underrepresented: “*To achieve care transformation, collaboration is essential, interdisciplinary and between departments”*. (ZGT 1) Within the strategic dimension, participants emphasised the importance of clear initiators or “champions” to drive innovation projects, highlighting a missing element of strategic leadership. Moreover, they repeatedly pointed to the influence of the external strategic and institutional context (e.g. health insurers, compliance, and regulation), which they felt was insufficiently captured in the current model: “*The external environment is an important factor in care”.* (MST 2).

#### Practical application of the OeHR model

Overall, participants considered the OeHR model a valuable framework for guiding care transformation, especially in supporting strategic decision-making and putting digital and hybrid care on the agenda. *“The model and the questionnaire give a clear direction for care transformation. If you do that at management levels, you can better determine where the focus should be”.* (MST 2) At the same time, the abstract nature of the model was viewed as a limitation, with a need for more concrete tools: *“We know with the OeHR what the problems are, but how do we actually tackle them?”*. (MST 2) The accompanying questionnaire was generally recognised as a practical instrument for assessing an organisation’s current position, provided it could be adapted to the specific context: *“Then you actually have to provide customisation, adapt per context”*.(ZGT 1) The model was seen as having potential to create space for dialogue in an environment where professionals are faced with time pressure and high. *“Many healthcare professionals are so busy with daily practice that they have little time to think about the impact of technological innovations”.* (MST 2)

## Discussion

### Reflecting on the existing OeHR

This study provides new insights into the applicability of the OeHR model in the Dutch hospital context. While the framework conceptualises organisational eHealth readiness as a multidimensional construct, the findings indicate that the five dimensions are highly interrelated and collectively reflect a single, integrated readiness factor. Cultural readiness, in particular, plays a pivotal role within this composite construct, suggesting that shared values, openness to change, and leadership support are central to shaping perceptions of organisational readiness for digital transformation. The other dimensions of OeHR also contribute to this overarching readiness, seemingly functioning as important contextual enablers whose impact is amplified when a supportive and adaptive organisational culture is present. These results highlight the need to consider both the individual dimensions and their interplay when assessing and fostering Organisational eHealth Readiness.

This study also examined stakeholders’ perceptions of the relevance, usefulness and areas for improvement of the OeHR model in Dutch hospitals. Insights from the focus groups confirmed the relevance of all five dimensions but revealed different priorities. Culture was consistently rated as most relevant, while structure was considered the least relevant. In addition, participants expressed disagreement about the current definition, content, and boundaries of culture. This may be in line with Greenhalgh et al. (2017), who position culture as a structural factor that can both promote and hinder implementation [24], which may be indicative of its complex and broad nature. Fowe (2021), in evaluating the CFIR framework, similarly noted that cultural preparedness depends not only on values and norms but also on leadership alignment and organisational agility [25,26]. The positive predictive value of cultural readiness found in this current study confirms that an organisational culture that is open to change, innovation and digital transformation is a crucial facilitator for eHealth adoption. According to Weiner’s Theory of Organisational Readiness for Change (2009), culture plays a central role in shaping both readiness for change and the effectiveness of change, the shared determination to pursue change, and collective confidence in the organisation’s ability to succeed [11]. From this perspective, cultural readiness is not just a behavioural factor, but an important mechanism through which members of the organisation mobilise the motivation and capacity necessary for transformation. This theoretical view reinforces the finding that culture acts as the main driver of perceived readiness for e-health and influences strategy, competencies, structure, and technology deployment. These findings suggest that cultural readiness cannot be viewed in isolation, as it is closely linked to other organisational factors and can be interpreted in different ways depending on the organisational perspective.

Focus group discussions and survey analyses together suggest that organisational eHealth readiness is best understood as an integrated construct, encompassing strategic, competence, structural, technological, and cultural dimensions. While technological infrastructure, strategic planning, and other structural and competence-related factors were considered important enablers in the focus groups, their influence on perceived readiness appears conditional on a supportive and adaptive organisational culture. Cultural readiness, reflecting shared values, openness to change, and leadership support, emerged as particularly central, strongly contributing to the composite readiness factor identified through PCA. These findings highlight that the five dimensions collectively shape perceptions of readiness, with cultural aspects playing a pivotal role in enabling and amplifying the impact of the other dimensions. This aligns with broader theoretical perspectives, which similarly emphasise that technology alone does not guarantee implementation success. Greenhalgh et al. (2004) describe technological readiness as a necessary but not sufficient condition for successful adoption [27]. In addition, Boonstra and Broekhuis (2010) emphasise that although structural readiness, such as policies and resources, provides essential preconditions, cultural acceptance and user commitment ultimately determine whether eHealth innovations take root in practice [28]. The current findings reinforce these insights: despite being viewed as important by participants, strategic and competence-related readiness did not show a significant independent contribution to overall organisational eHealth readiness in the quantitative analyses. This contrasts with previous studies in which digital literacy, training, and leadership are often cited as key conditions for digital transformation [29]. A possible explanation for the lack of a significant independent contribution of strategic and competence eHealth readiness in this study may be related to methodological and contextual factors. First, the relatively small sample size (N = 22) likely reduced the statistical power of the regression analyses, limiting the ability to detect unique effects of individual predictors. Second, substantial multicollinearity between the OeHR dimensions may have masked their independent contributions. However, looking beyond methodological constraints, previous research also suggests that strategic and competence-related readiness may exert influence only when more fundamental organisational conditions, such as a supportive digital culture, effective leadership, and sufficient technological capacity, are already in place [18]. This interpretation is consistent with the qualitative findings of this study, which underline that cultural readiness serves as a critical integrator that shapes how other readiness dimensions translate into perceived organisational preparedness for eHealth implementation.

#### Overall applicability of the OeHR model

The combined findings suggest that the OeHR model can play a meaningful role in supporting care transformation by encouraging organisations to broaden their perspective and align strategic, tactical, and operational considerations more effectively [18]. Although originally developed as a diagnostic framework, participants in this study highlighted the need for complementary practical guidance to bridge the gap between identifying challenges and implementing concrete actions [30,31].

The OeHR questionnaire was perceived as useful for mapping an organisation’s current position. It was also seen as a valuable tool to foster dialogue in a sector that is often constrained by high time pressure and workload, which can limit opportunities for structured reflection.

At the same time, the quantitative results indicate that when respondents assess their organisation’s overall eHealth readiness, they primarily draw on perceptions of cultural readiness. This suggests that perceived organisational readiness is currently understood largely through a cultural lens. In contrast, the OeHR framework offers a broader, multidimensional perspective that also encompasses strategic, competence-related, structural, and technological aspects of readiness.

To enhance practical applicability, adaptive and context-sensitive tools may better guide decision-making by accounting for organisational, infrastructural, and policy-related conditions [32]. Evidence on organisational readiness for change further shows that shared commitment, motivation, and resource availability are critical for successful implementation [33]. Validated readiness self-assessments can help organisations identify strengths and gaps and take targeted measures in terms of infrastructure, training, and governance, thereby accelerating adoption and improving quality [13].

#### Strengths, limitations and future research

Strength of the present study is the inclusion of participants from different regions of The Netherlands, the use of a validated questionnaire and the mixed-methods approach to combine data with additional in-depth information. When interpreting the data, the relatively small sample size and the presence of multicollinearity that limit the statistical validity of the regression outcomes and constrain the generalisability of the findings to the broader hospital context need to be considered. Future research with larger samples is needed to validate these relationships and further clarify the distinct and shared contributions of the different readiness dimensions.

Building on the findings of this study, future refinement of the OeHR model should prioritise enhancing conceptual clarity and delineating more precise boundaries between the strategic, structural, and cultural dimensions. Strengthening definitional precision would reduce conceptual overlap and improve the model’s internal coherence. In addition, the model could be enriched by integrating more dynamic aspects, such as behavioural flexibility within culture, organisational agility and collaboration within structure, and the influence of institutional environments and innovation champions within strategy. Such refinements would not only increase the theoretical robustness of the model but also enhance its practical diagnostic value for healthcare organisations seeking to assess and improve their readiness for digital transformation.

It remains unclear whether these dimensions need to reach a certain level before readiness becomes apparent, or whether they are simply less noticeable at present. Future studies should therefore investigate the extent to which each dimension contributes to organisational readiness once more fully developed, and whether the central role of cultural readiness persists when all dimensions are adequately represented. Such longitudinal and comparative approaches could yield a more dynamic understanding of how the various facets of organisational eHealth readiness evolve and interact over time.

Beyond refining the existing dimensions, future research could also explore the inclusion of complementary forms of readiness that broaden the organisational focus of the current model. The findings tentatively suggest the relevance of personal eHealth readiness (individual skills and motivation to engage with digital care), operational eHealth readiness (the feasibility and organisational support for implementing eHealth solutions), and societal eHealth readiness (external conditions such as legal, regulatory, and financial factors). Although these aspects emerged only sporadically in this study, they highlight potential systemic and individual layers that may influence an organisation’s capacity for digital transformation. Exploring how these additional forms of readiness interact with organisational factors could provide valuable directions for future theoretical development and empirical validation of the OeHR framework.

## Conclusion

This study set out to examine how stakeholders in Dutch hospitals perceive and assess the OeHR model, and to analyse the quantitative relationship between its underlying components and perceived organisational eHealth readiness. This study suggests that the Organisational eHealth Readiness (OeHR) model may offer a useful framework for assessing and guiding care transformation in the context of top clinical hospitals. Through its mixed-methods design, the study provides exploratory insights into how the underlying dimensions of the model may contribute to organisational readiness for eHealth. Cultural eHealth Readiness is widely recognised by professionals as a crucial factor, yet its definition requires further refinement and definition. In contrast, strategic, competence, structural and technological eHealth readiness appear to function primarily as a contextual enabler or boundary condition; they constitute essential prerequisites for care transformation, [2] yet do not in themselves strengthen or advance the overall level of organisational eHealth readiness. The study identified three main eHealth readiness dimensions, personal, operational, and societal along with factors like leadership, change readiness, collaboration, user experience, regulation, capacity, flexibility, and external context missing from the current OeHR model. In addition, participants emphasised the organisation’s ability to manage and absorb change as a crucial factor influencing how effectively eHealth initiatives can be implemented and sustained. Incorporating these items and dimensions in future iterations could enhance the comprehensiveness and practical applicability of the OeHR framework.

## Data Availability

Data can be accessed in DANS data repository via: https://doi.org/10.17026/LS/BNPH87

## Supporting Information

**S1 Keywords per dimension**

